# Progression of fiber bundle damage in amnestic Alzheimer’s disease and LATE: a 2-year fixel-based study

**DOI:** 10.1101/2025.09.05.25333680

**Authors:** Aurélie Lebrun, Yann Leprince, Pauline Olivieri, Martin Moussion, Camille Noiray, Michel Bottlaender, Marie Sarazin, Julien Lagarde

## Abstract

Alzheimer’s disease (AD) and limbic-predominant age-related TDP-43 encephalopathy (LATE) are two neurodegenerative diseases underpinned by distinct proteinopathies, which share a similar initial amnestic clinical phenotype but have a different clinical course. Some pathophysiological models suggest a role for white matter (WM) fiber bundles in disease progression, but they remain discussed, as does their applicability to LATE. Using longitudinal fixel-based analysis, we investigated the progression of WM fiber bundle alterations over two years in early amnestic AD (n=16; clinical-biological diagnosis based on CSF biomarkers and amyloid/tau-PET imaging), probable LATE (n=12; diagnosis based on recently published diagnostic criteria), and amyloid-negative controls (n=15). We explored the associations between baseline and longitudinal WM fiber bundle alterations and (i) cognitive/functional decline in both patient groups, and (ii) baseline amyloid and tau load in patients with AD. In both AD and LATE, we found a 2-year progression of WM fiber bundle alterations in temporo-parietal and temporo-frontal tracts, most of which were altered at baseline. We also found a differential 2-year progression of the alterations according to pathology, affecting temporal and limbic tracts in AD, and the ventral section of the superior longitudinal fasciculus in LATE. Low metrics within these fiber bundles at baseline were associated with a more rapid cognitive decline. We found no association between WM fiber bundle alterations and baseline tau-PET and only weak associations with amyloid-PET. These results suggest a functional role for WM fiber bundle alterations in the progression of cognitive impairment, making this parameter a potential predictive marker of cognitive decline.

## Background

Progressive limbic/episodic amnestic syndrome associated with hippocampal atrophy is often caused by Alzheimer’s disease (AD), but other causes, such as Limbic-predominant age-related TDP-43 encephalopathy (LATE), are not uncommon.^1^ LATE is characterized by the aggregation and accumulation of abnormal TDP-43 with or without coexisting hippocampal sclerosis. During aging, LATE is often mistaken as early AD due to its initial amnestic phenotype.^1,2^ In the absence of *in vivo* biomarkers for TDP-43, LATE can only be diagnosed with certainty at autopsy. LATE clinical diagnosis may be presumed in individuals with isolated amnestic syndrome and medial temporal lobe atrophy when AD biomarkers (CSF, molecular PET imaging) are negative.^1,3,4^ With the emergence of therapeutic strategies targeting AD pathophysiology, it is important to better understand how LATE differs from early AD.

Although AD and LATE are mainly considered grey matter diseases, alterations in white matter (WM) have recently gained increasing attention, and may also contribute to their clinical expression.^5–7^ Diffusion MRI is considered the method of choice for studying WM alterations *in vivo*, with most studies relying on diffusion tensor imaging (DTI). In AD, cross-sectional studies have shown WM damage in a number of tracts, particularly in the temporal lobe.^8,9^ Longitudinal studies have also been performed, reporting the alteration of several WM tracts but the results remain disparate.^10–16^ In LATE, only few recent studies have been conducted to explore WM alterations cross-sectionally,^17-21^ and no previous longitudinal study has been reported.

Fixel-based analysis is a recently developed method that allows studying WM at the level of each fiber population within a voxel, which overcomes the major limitation of most diffusion MRI models in the presence of fiber crossings.^22,23^ It provides metrics at both the microstructure (fiber density) and the macrostructure (fiber bundle atrophy) levels. This method has been used to study WM alterations cross-sectionally in AD.^24-26^ In a previous study, we showed alterations of WM fiber bundles in AD and probable LATE, with partly distinct topographical patterns between the two diseases.^27^ No fixel-based study has yet investigated the longitudinal evolution of WM fiber bundle alterations in both AD and LATE in comparison with healthy controls.

In this study, for the first time to the best of our knowledge, we explored the longitudinal evolution of WM fiber bundle alterations in AD and probable LATE (defined by strict clinical-biological criteria) over two years in comparison to amyloid-negative controls using fixel-based analysis. We also investigated the associations between baseline and longitudinal WM alterations and cognitive/functional decline over two years in each group of patients. Finally, in the AD group, we analyzed the relationship between baseline amyloid and tau load and WM fiber bundle damage. We hypothesized a progression of WM fiber bundle alterations over time, possibly exhibiting partially distinct topographical patterns in AD and LATE, and that these WM fiber bundle alterations may be associated with cognitive decline.

## Methods

### Study participants

This study included 43 participants from the prospective SHATAU7/IMATAU cohort (EudraCT: 2015-000257-20).^27^ All subjects provided written informed consent. The study was approved by a French Ethics Committee (CPP Île-de-France VI). All participants underwent complete clinical and neuropsychological assessment, 3-tesla brain MRI, lumbar puncture for AD CSF biomarkers (except for controls), amyloid PET imaging using [^11^C]Pittsburg compound B ([^11^C]PIB) and tau PET imaging using [^18^F]-Flortaucipir at inclusion (session 1). Participants were then followed up annually for two years with the same clinical and neuropsychological assessment, and underwent a second 3-tesla brain MRI at the last visit (session 2).

Patients with early Alzheimer’s disease (n=16) were included according to the following criteria: (i) progressive amnestic syndrome of the hippocampal type, (ii) Clinical Dementia Rating (CDR) = 0.5 or 1; (iii) CSF biomarkers suggestive of AD (phosphorylated-tau/amyloid-beta 42 > 0.08); and (iv) positive amyloid and tau PET imaging.

Patients with probable LATE (n=12) were included according to the following criteria: (i) progressive amnestic syndrome of the hippocampal type; (ii) CDR = 0.5 or 1; (iii) CSF biomarkers not suggestive of AD; (iv) negative amyloid and/or tau PET imaging; (v) no extrapyramidal signs or other neurological signs suggestive of Parkinson’s disease, progressive supranuclear palsy, corticobasal degeneration, frontotemporal dementia, dementia with Lewy bodies; (vi) no vascular lesion on MRI that could account for amnesia, and (vii) absence of neurological diagnoses other than probable LATE after two years of clinical follow-up, including a second brain MRI for all subjects and a second tau PET imaging when available (n=8/12). Note that two patients had slightly positive amyloid PET imaging despite normal CSF biomarkers and negative baseline and longitudinal tau PET imaging. As proposed in the recently published LATE clinical criteria,^4^ they were considered patients with possible LATE and amyloid co-pathology, and included in the study. To facilitate reading, we will refer to probable/possible LATE as LATE in the remainder of this article.

Healthy controls (n=15) were included according to the following criteria: at both sessions (i) Mini-Mental State Examination (MMSE) score ≥ 27/30 and normal neuropsychological assessment; (ii) CDR = 0; (iii) no memory complaints; (iv) negative amyloid and tau PET imaging.

### MRI acquisition and preprocessing

MRI were all acquired at the Paris Brain Institute using a 3-tesla Siemens Magnetom Prisma scanner with a 64-channel coil, including for each visit, T1-weighted images and diffusion-weighted images (DWI). Each DWI comprised three shells of diffusion with b-values of 200, 1700, 4200 s/mm^2^ and 60 directions per shell, and three volumes without diffusion weighting (b = 0 s/mm ^2^). Acquisition parameters and preprocessing steps of the images have been published previously^27^ and are described in Supplementary file 1. Briefly, DWI were denoised and corrected for head motion, and susceptibility and eddy current induced distortions.^28,29^ Then, fiber orientation distributions (FODs) were obtained at each voxel using multi-shell multi-tissue spherical deconvolution,^30^ and joint bias field correction and global intensity normalization were applied.

### Longitudinal fixel-based analysis

All the following steps were performed using MRtrix3 commands (version 3.0.3).^31^ A pseudo-code of the pipeline used is available in Supplementary file 2. To perform longitudinal fixel-based analysis, we used the common inter-subject FOD template that we previously computed^27^ using the FODs at session 1 of 8 patients with AD, 8 patients with LATE, and 16 controls (see Supplementary file 3 for details on the choice of template).

We then registered each subjects’ FODs on the common template, using a two-step registration method to maximize sensitivity to longitudinal effects.^32^ For each subject, FODs of both sessions were first non-linearly co-registered, transformed to the midway space between the two sessions, and averaged to obtain an intra-subject FOD average (step 1). This intra-subject FOD average was then non-linearly registered to the common inter-subject template (step 2). The two deformation fields resulting from the two registration steps were then composed to generate a single deformation field per session, which was reinjected in the usual fixel-based analysis pipeline to transform the native FODs to the common inter-subject template without performing multiple interpolations. For the second registration step we decreased the default values of the regularization parameters, in order to optimize the registration of these images with different levels of atrophy, e.g. ventricular size.

We then obtained a brain mask of the template by intersecting all subjects’ masks in the template space. We removed from the mask the areas where the confusion between WM and CSF compartments among subjects was too extensive, as previously described.^27^ This enabled to minimize spurious results due to remaining periventricular registration inaccuracies.

Finally, we segmented the registered FODs into fixels and we calculated the fixel metrics in each fixel of each image: fiber density (FD), fiber bundle cross-section (FC), and fiber density and cross-section (FDC = FD x FC). While FD can be interpreted as a relative measure of axonal density, FC measures fiber bundle macroscopic atrophy. FDC is a combined measure of FD and FC and captures fiber loss due to either reduction in fiber density and/or macroscopic atrophy. FC maps were log-transformed.^22^

In order to investigate FD, FC and FDC changes between sessions 1 and 2 while accounting for variation in inter-session interval, we computed an annualized rate of change R of FD, log(FC), and FDC as follows:

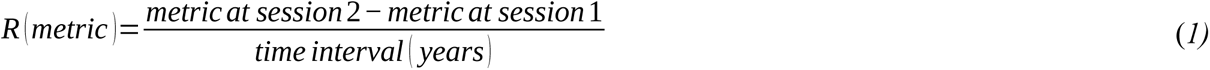

### Whole-brain fixel-based analyses

We first performed exploratory whole-brain fixel-based analyses to investigate the possible decrease in fixel metrics over two years at the level of each fixel within each participant group and between groups. To do so, we used a whole-brain tractogram generated on the common template using probabilistic tractography with twenty million fibers that was subsequently filtered to two million fibers.^33^ We then computed the connectivity matrix between fixels of the template and applied subsequent connectivity-based smoothing.^34^

### PET acquisition and processing

Participants underwent [^11^C]PIB PET imaging (missing data for one AD patient and one control) and [^18^F]-Flortaucipir PET imaging (missing data for two AD patients and one control) at inclusion. All PET examinations were performed at Service Hospitalier Frédéric Joliot (Orsay, CEA) on a High-Resolution Research Tomograph (CTI/Siemens Molecular Imaging). PET acquisitions were performed 40 to 60 min after injection of 332 ± 60.8 MBq of [^11^C]PIB, and 80–100 min after injection of 376.2 ± 20.6 MBq of [^18^F]-Flortaucipir. Details on PET processing have been previously published^2^ and are described in Supplementary file 1.

### Definition of the cognitive outcomes

Based on our previous work,^35^ in addition to the MMSE score as a measure of global cognitive assessment and to the CDR sum of boxes score as a measure of functional status, we defined a verbal episodic memory score, a parietal score, and an executive score (see Supplementary file 1 for details). For each of these composite scores, we computed the annualized rate of change R (Equation 1) between sessions 1 and 2.

### Statistical analyses

#### Whole-brain fixel-based analyses

For each participant group, we first performed within-group whole-brain fixel-based analyses to test, at the level of each fixel, if the annualized rates of change in FD, FC and FDC (R(FD), R(log(FC)) and R(FDC)), were significantly lower than 0. We did not use covariates for these longitudinal analyses.

We then performed a between-group whole-brain fixel-based analysis to compare the R(FD), R(log(FC)), and R(FDC) between (i) AD and controls, and (ii) LATE and controls. We computed statistical tests for each metric at each fixel, using a general linear model (GLM) including age at baseline and sex as covariates.

For both analyses, statistical inferences were performed using connectivity-based fixel enhancement^34^ using non-parametric permutations and default parameters except for within-group analyses, where the nature of the errors for shuffling was set to independent and symmetric. The significance of the results was assessed using family-wise error correction with a type I error rate of 5%.

#### Tract-based analyses

Based on the whole-brain fixel-based analyses, we identified 11 relevant fiber bundles that we reconstructed on the common FOD template. Additionally, in a previous cross-sectional study performed on slightly larger participant groups from the same cohort,^27^ we highlighted the alteration of 13 other WM tracts in either patient group. We reconstructed all these tracts on the common FOD template using TractSeg^36^ or probabilistic maps of presence of the tract extremity regions, as previously described.^27^ Anatomical descriptions of the extremity regions of each tract are summarized in Supplementary file 4.

For the 24 reconstructed tracts, we performed tract-based analyses on the FDC values obtained at session 1 and on the R(FDC) in both patient groups compared to controls. We calculated, for each tract, the mean baseline FDC and the mean R(FDC) for the whole tract by taking the FDC average or the average of each R(FDC) over all fixels associated with the tract, weighted by streamline density. We then performed statistical tests to compare the mean baseline FDC and the mean R(FDC) between (i) AD and controls, and (ii) LATE and controls, using a GLM with age, sex, and intracranial volume (the latter only for the mean baseline FDC) as covariates.

For both analyses, the significance of the results was assessed with one-sided t tests, performing false discovery rate correction among the 48 tests performed per analysis with a type I error rate of 5%.

#### Association between white matter alterations and cognitive trajectories

The analyses were performed within each patient group, for each tract that showed a significant decline in FDC over time compared to controls. In a predictive study, we first explored associations between mean baseline FDC and cognitive/functional decline (R(MMSE), R(CDR sum of boxes), R(parietal score), and R(executive score)) using age, sex, and intracranial volume as covariates. We then analyzed the correlations between the mean R(FDC) and the cognitive/functional decline, using age at baseline and sex as covariates. Significance of the results was assessed with a type I error rate of 5%. It should be noted that for one patient with AD, the memory score at two years was not measurable due to floor effect. In addition, as delayed recall was not measurable at two years in most patients, we did not test for the associations between WM fiber bundle alterations and memory cognitive trajectory.

#### Association between white matter fiber bundle alterations and PET imaging in the AD group

The analyses were performed in patients with AD, for each tract that showed a significant decline of FDC over time compared to controls. We first analyzed the associations between amyloid (global cortical index and Standardized Uptake Value ratio (SUVr) in the precunei) or tau (global cortical index and SUVr in the temporal meta-volume of interest) deposition and the FDC values at inclusion using age, sex, and intracranial volume as covariates. We then investigated the association between amyloid or tau SUVr at inclusion and R(FDC) in the same tracts using age and sex as covariates. Significance of the results was assessed with a type I error rate of 5%.

## Results

### Characteristics of the participants

Demographic and clinical data of participants at both sessions are summarized in Table 1. At inclusion, patients with AD and patients with LATE had similar clinical phenotypes. Patients with LATE were older and exhibited greater hippocampal atrophy than patients with AD. The annualized rates of change R showed a higher decline in both the memory and the parietal composite scores in patients with AD than in the other groups (mean R(memory score) = -8.0, -2.3, 1.3; mean R(parietal score) = -6.1, 0.1, 0.4 for AD, LATE, and controls respectively). Finally, each group of participants exhibited different reductions in normalized hippocampal volume with a higher rate of change for patients with AD (-0.008 for AD, -0.004 for LATE, and -0.001 for controls).

**Table 1.** Demographic, clinical, and imaging data at sessions 1 and 2.

|  | Session 1 |  |  |  | Session 2 |  |  |  | Test for between-group differences in rates of change R |
| --- | --- | --- | --- | --- | --- | --- | --- | --- | --- |
|  | Alzheimer's disease (n=16) | Probable LATE (n=12) | Controls (n=15) | Test for between-group differences | Alzheimer's disease (n=16) | Probable LATE (n=12) | Controls (n=15) | Test for between-group differences |  |
| **Demographics** |  |  |  |  |  |  |  |  |  |
| Age (SD) | 72.6 (6.0) | 78.8 (4.9)* | 68.3 (3.6) | $F = 13.9, P < 0.001$ | 74.6 (6.0) | 80.8 (5.0)* | 70.2 (3.5) | $F = 14.1, P < 0.001$ | $F = 2.7, P = 0.082$ |
| Males (%) | 7 (44) | 8 (67)* | 5 (38) | $\chi^2 = 9.1, P = 0.011$ | - | - | - | - | - |
| Years of education (SD) | 14.8 (4.4) | 14.8 (5.3) | 14.7 (3.0) | $F = 0.0, P = 0.996$ | - | - | - | - | - |
| **Medication** |  |  |  |  |  |  |  |  |  |
| Cholinesterase inhibitors (n) | 11 | 1 | 0 | - | Introduced in 1, discontinued in 2 | - | - | - | - |
| Memantine (n) | 1 | 0 | 0 | - | - | - | - | - | - |
| **Functional status** |  |  |  |  |  |  |  |  |  |
| CDR (0/0.5/1/2) | 0/15/1/0 | 0/9/3/0 | 15/0/0/0 | - | 0/4/8/4 | 0/6/5/1 | 15/0/0/0 | - | - |
| CDR sum of boxes (SD) | 3.7 (1.8) | 3.8 (2.3) | 0.0 (0.0)* | $F = 24.6, P < 0.001$ | 6.4 (2.9) | 5.8 (2.8) | 0.0 (0.0)*† | $F = 32.5, P < 0.001$ | $F = 11.6, P < 0.001$ |
| **Cognitive scores** |  |  |  |  |  |  |  |  |  |
| MMSE /30 (SD) | 24.3 (3.2) | 25.0 (2.9) | 28.9 (0.8)* | $F = 13.4, P < 0.001$ | 20.3 (4.9) | 22.9 (3.5) | 29.7 (0.6)*† | $F = 26.9, P < 0.001$ | $F = 12.8, P < 0.001$ |
| Memory score: FCSRT /128 (SD) | 47.8 (25.2) | 49.0 (22.1) | 110.1 (7.3)* | $F = 44.3, P < 0.001$ | 31.1 (30.8)† | 43.7 (21.4) | 112.4 (5.9)* | $F = 54.0, P < 0.001$ | $F = 9.3, P < 0.001$ |
| Parietal score: Naming + praxis + Rey figure copy /188 (SD) | 180.0 (3.5) | 181.4 (3.5) | 185.9 (1.6)* | $F = 15.3, P < 0.001$ | 167.3 (21.6)*† | 181.5 (3.6) | 186.7 (1.8) | $F = 8.2, P = 0.001$ | $F = 5.8, P = 0.006$ |
| Executive score: Digit spans + letter fluency 2 min + WAIS similarities (SD) | 49.3 (11.7) | 51.6 (6.8) | 62.7 (9.0)* | $F = 7.7, P = 0.001$ | 48.0 (17.7) | 49.2 (11.0) | 65.0 (11.4)* | $F = 6.3, P = 0.004$ | $F = 0.8, P = 0.45$ |
| **Molecular PET imaging** |  |  |  |  |  |  |  |  |  |
| Amyloid PET SUVR, GCI (SD)<sup>a</sup> | 2.88 (0.69)* | 1.38 (0.27) | 1.28 (0.10) | $F = 53.9, P < 0.001$ | - | - | - | - | - |
| Tau PET SUVR in a temporal meta-VOI (SD)<sup>b</sup> | 2.11 (0.83)* | 1.22 (0.09) | 1.23 (0.13) | $F = 13.3, P < 0.001$ | 2.27 (0.63)* | 1.25 (0.10) | 1.26 (0.11) | $F = 22.3, P < 0.001$ | $F = 3.6, P = 0.040$ |
| **MRI** |  |  |  |  |  |  |  |  |  |
| Time interval in years between session 1 and session 2 (SD) | - | - | - | - | 2.05 (0.12) | 2.19 (0.34) | 1.95 (0.36) | $F = 2.2, P = 0.123$ | - |
| Intracranial volume (SD) | 1566 (160) | 1685 (134) | 1585 (108) | $F = 2.7, P = 0.080$ | - | - | - | - | - |
| Normalized HV (SD)<sup>c</sup> | 0.205 (0.027)* | 0.170 (0.039)* | 0.246 (0.019)* | $F = 21.2, P < 0.001$ | 0.189 (0.029)† | 0.162 (0.040)† | 0.244 (0.019)*† | $F = 25.4, P < 0.001$ | $F = 25.5, P < 0.001$ |
| Fazekas score (0/1/2/3) | 10/4/1/1 | 5/5/1/1 | 9/5/1/0 | - | 8/6/1/1 | 3/5/3/1 | 9/5/1/0 | - | - |

**Notes:** Mean values (standard deviation) or numbers (%) are presented for each data at visit 1 and visit 2.
For each visit, statistics between groups are reported with P values from one-way between-groups ANOVA for continuous variables and chi-squared tests for categorical variables. Post hoc Tukey honest significance difference tests were then performed to compare each group to the others.
Statistics to compare between groups the 2-year evolutions of the data between visit 1 and 2 (computed as annualized rates of change R, see equation 1) with one-way between-groups ANOVA are reported in the last column. Post hoc Tukey honest significance difference tests were then performed to compare each group to the others.
Abbreviations: CDR, Clinical Dementia Rating; FCSRT, Free and Cued Selective Reminding Test; GCI, Global Cortical Index; HV, hippocampal volume (normalized to intracranial volume); MMSE, Mini Mental State Examination; SD, standard deviation; SUVR, standardized uptake value ratio; VOI, volume of interest; WAIS, Wechsler Adult Intelligence Scale.
\*P < 0.05 versus the two other groups in the comparisons at visit 1 or 2 (assessed with post-hoc Tukey honest significance test)
†P < 0.05 versus the two other groups in the comparisons of the 2-year rates of change (assessed with post-hoc Tukey honest significance test)
<sup>a</sup>Missing data for 1 patient with AD and 1 control
<sup>b</sup>Missing data for 2 patients with AD and 1 control at visit 1, and for 3 patients with AD, 4 patients with probable LATE and 3 controls at visit 2
<sup>c</sup>One patient with AD was excluded because of insufficient T1-weighted image quality

### Whole-brain fixel-based analyses

We first tested, within each group, whether FD, FC, or FDC decreased significantly between the two visits. We found significant results for FC in both patient groups as shown in Figure 1. Patients with AD exhibited a significant decrease in FC in the inferior longitudinal, middle longitudinal, arcuate, and superior longitudinal III fasciculi, in the dorsal cingulum, and in callosal fibers connecting the precunei. We also found significant decrease in FDC in AD in areas that overlapped with the results obtained for FC. Patients with LATE exhibited significant decrease in FC in a temporal region that can be attributed to both the arcuate fasciculus and/or the middle longitudinal fasciculus. We found no significant decrease in FD in either group of patients. There was no significant reduction in FD, FC, or FDC in the control group.

**Figure 1.**
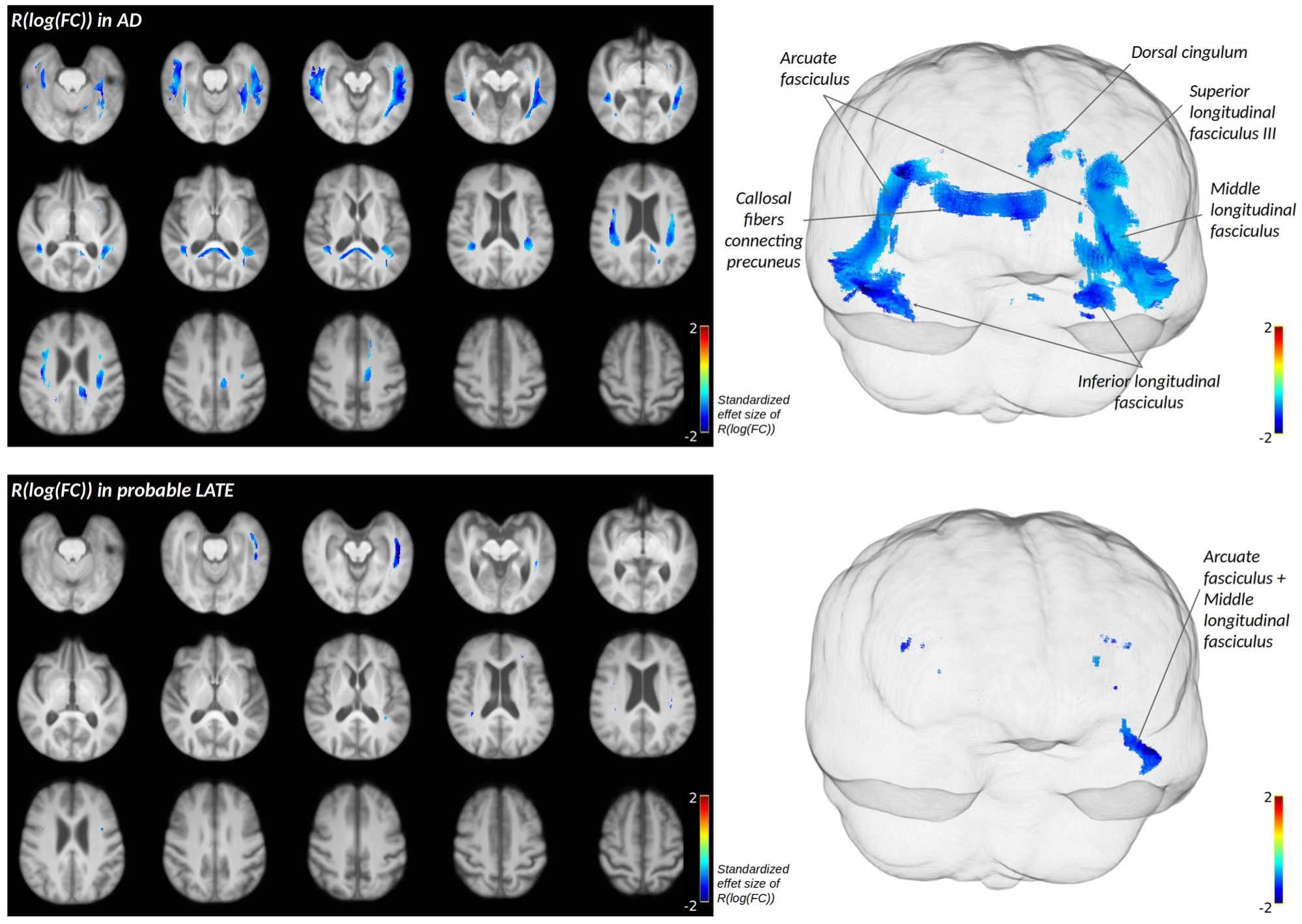
Within-group whole-brain fixel-based analyses, within patients with AD (top), and patients with probable LATE (down). Fixels exhibiting significant decrease in FC within each group (test: R(log(FC)) < 0, no covariates, p < 0.05 after family-wise error correction) are represented as cropped fibers from the template-derived tractogram and are colored by standardized effect sizes. Results are displayed across axial slices of a mean T1-weighted image across subjects on the left, and through a glass brain representation on the right.

We then performed a between-group analysis in each patient group compared to controls, while controlling for age and sex. We found significant differences only for FC in the arcuate and middle longitudinal fasciculi in AD, and in the superior longitudinal fasciculus III in LATE (Figure 2).

**Figure 2.**
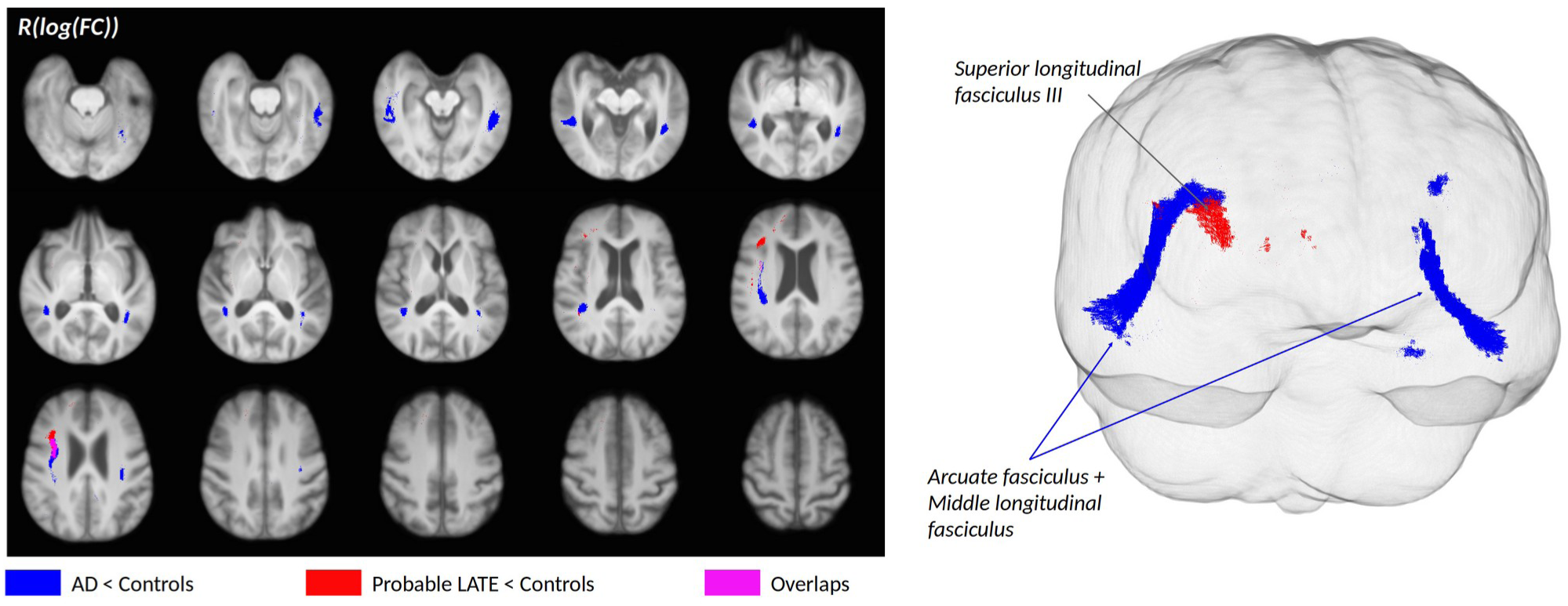
Between-group whole-brain fixel-based analysis: decrease in FC is stronger in patients with AD (blue) and in patients with probable LATE (red) compared to healthy controls. Significant results (test: R(log(FC_patients)) < R(log(FC_controls)), covariates: age and sex, p < 0.05 after family-wise error correction) are represented as cropped fibers from the template-derived tractogram in blue for the comparison between patients with AD and controls and in red for the comparison between patients with probable LATE and controls. The pink shade corresponds to the overlap areas between the results obtained for each comparison. Results are displayed across axial slices of a mean T1-weighted image across subjects on the left, and through a glass brain representation on the right.

### Tract-based analyses

Figure 3 shows the results obtained for the comparison of the mean baseline FDC between each patient group and controls in the 24 tracts that were reconstructed. Baseline FDC was lower in patients with AD compared to controls in almost all tested tracts except some portions of the corpus callosum and the ventral cingulum. Baseline FDC was lower in patients with LATE compared to controls in multiple portions of the corpus callosum, in the cerebello-thalamo-cortical tract, and in the uncinate, superior longitudinal III, and middle longitudinal fasciculi.

**Figure 3.**
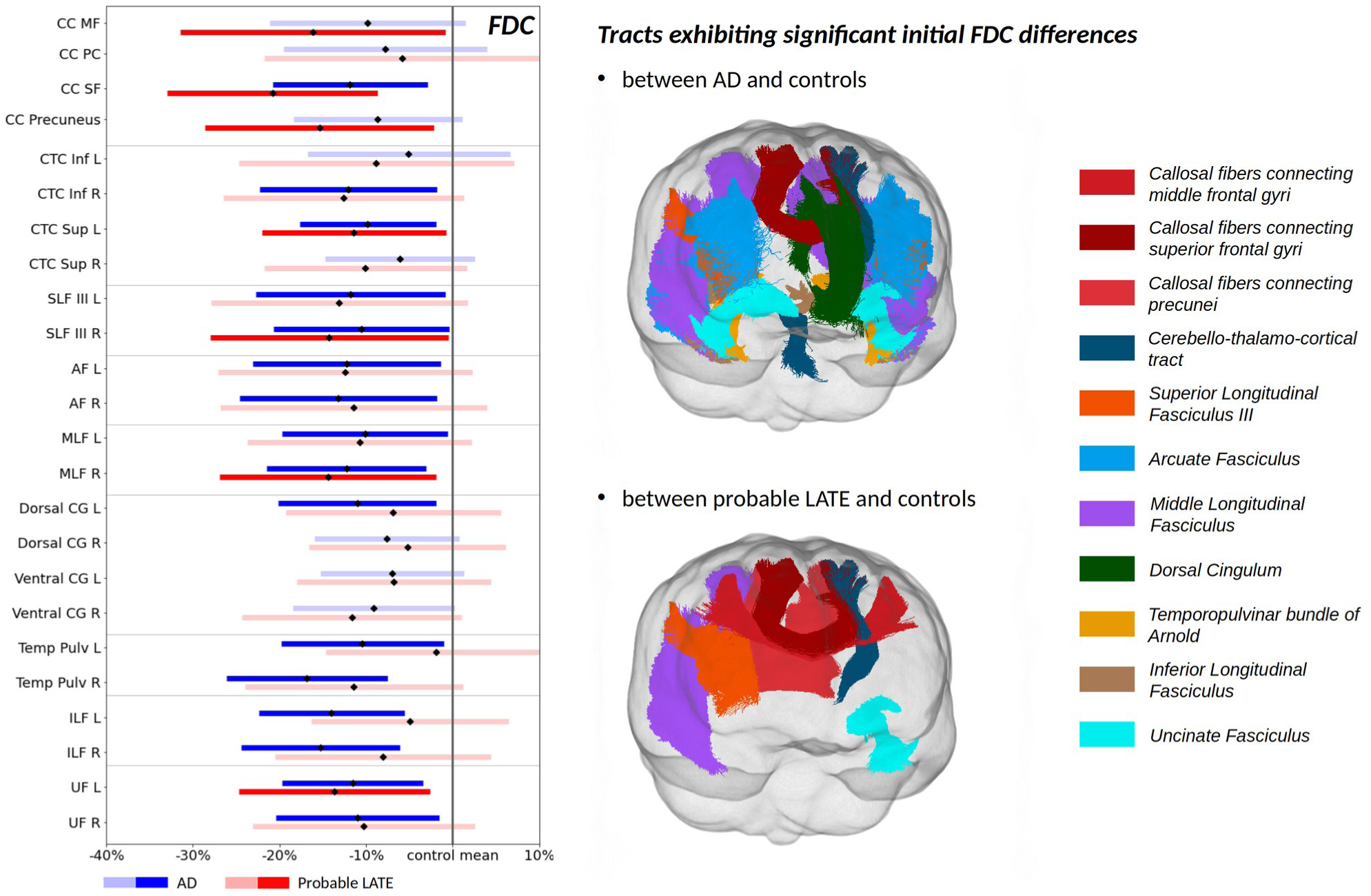
Tract-of-interest analyses comparing, in each tract, FDC values at baseline in patients with AD and in patients with probable LATE to that in controls. For each tract, the mean FDC value in patients with reference to controls (((FDC(patients) - FDC(controls))/FDC(controls)), is represented with diamonds and 95% confidence intervals with lines. Results are controlled for age, sex, and intracranial volume. For each tract, the blue line (first line) represents the comparison between patients with AD and controls, and the red line (second line) represents the comparison between patients with probable LATE and controls. Saturated colors highlight significant differences between patients and controls (p < 0.05 after false discovery rate correction). Tracts that exhibited significant results are displayed on the right of the figure. AF, arcuate fasciculus; CC, corpus callosum; CG, cingulum; CTC, cerebello-thalamo-cortical tract; ILF; inferior longitudinal fasciculus; Inf, inferior; L, left; MLF, middle longitudinal fasciculus; MF, middle frontal gyrus; PC, precentral gyrus; R, right; SF, superior frontal gyrus; SLF, superior longitudinal fasciculus; Temp Pulv, temporopulvinar bundle of Arnold; UF, uncinate fasciculus.

Figure 4 shows the results obtained for the comparison of the mean R(FDC) between each patient group and controls. The decrease in FDC was significantly stronger in patients with AD compared to controls in the ventral and dorsal cingulum, in the temporopulvinar bundle of Arnold, in the inferior and middle longitudinal fasciculi, and in the arcuate fasciculus. The decrease in FDC was significantly stronger in patients with LATE compared to controls in the middle and superior longitudinal III fasciculi, and in the arcuate fasciculus.

**Figure 4.**
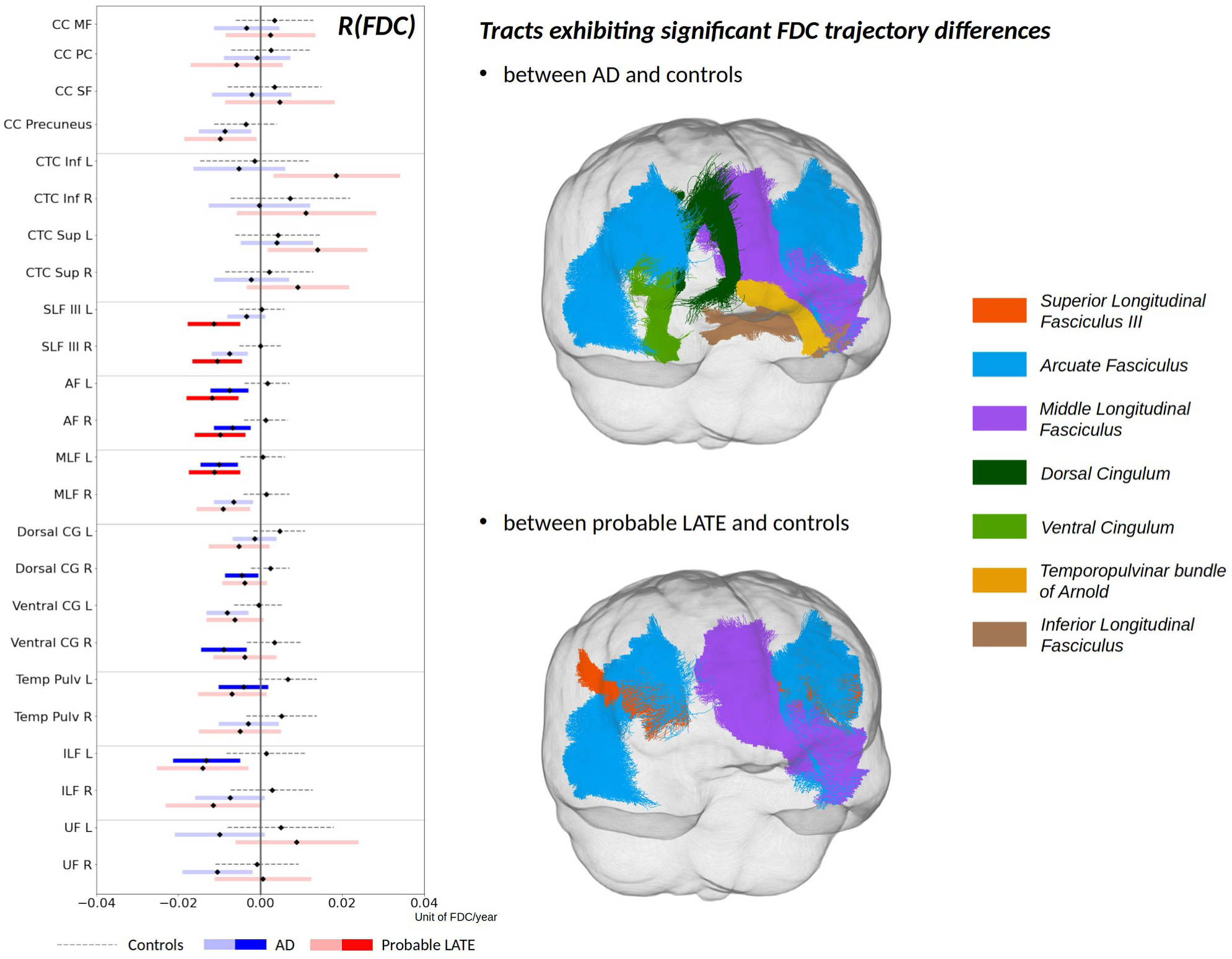
Tract-of-interest analyses comparing, in each tract, FDC annualized rate of change (R(FDC)) in patients with AD and in patients with probable LATE to that in controls. For each tract, the mean FDC annualized rate of change (R(FDC)) in each group of subjects is represented with diamonds and 95% confidence intervals with lines. Results are controlled for age and sex. For each tract, the dashed gray line represents the R(FDC) of controls, the solid blue line (first solid line) represents the R(FDC) of patients with AD, and the solid red line (second solid line) represents the R(FDC) of patients with probable LATE. Saturated colors highlight significant differences in R(FDC) between patients and controls (p < 0.05 after false discovery rate correction). Tracts that exhibited significant results are displayed on the right of the figure. AF, arcuate fasciculus; CC, corpus callosum; CG, cingulum; CTC, cerebello-thalamo-cortical tract; ILF; inferior longitudinal fasciculus; Inf, inferior; L, left; MLF, middle longitudinal fasciculus; MF, middle frontal gyrus; PC, precentral gyrus; R, right; SF, superior frontal gyrus; SLF, superior longitudinal fasciculus; Temp Pulv, temporopulvinar bundle of Arnold; UF, uncinate fasciculus.

### Association between white matter alterations and cognitive trajectories

Within the AD group, we found that (i) lower baseline FDC values in the arcuate fasciculi, the middle longitudinal fasciculi, the ventral cingulum bundles and the temporopulvinar bundles of Arnold were associated with stronger cognitive decline in parietal score and stronger functional decline; (ii) lower baseline FDC values in the dorsal cingulum bundles were associated with stronger decline in parietal score. Within the LATE group, lower baseline FDC values in the superior longitudinal fasciculi III, the middle longitudinal fasciculi and the arcuate fasciculi were associated with stronger decline in parietal score (Figure 5A).

**Figure 5.**
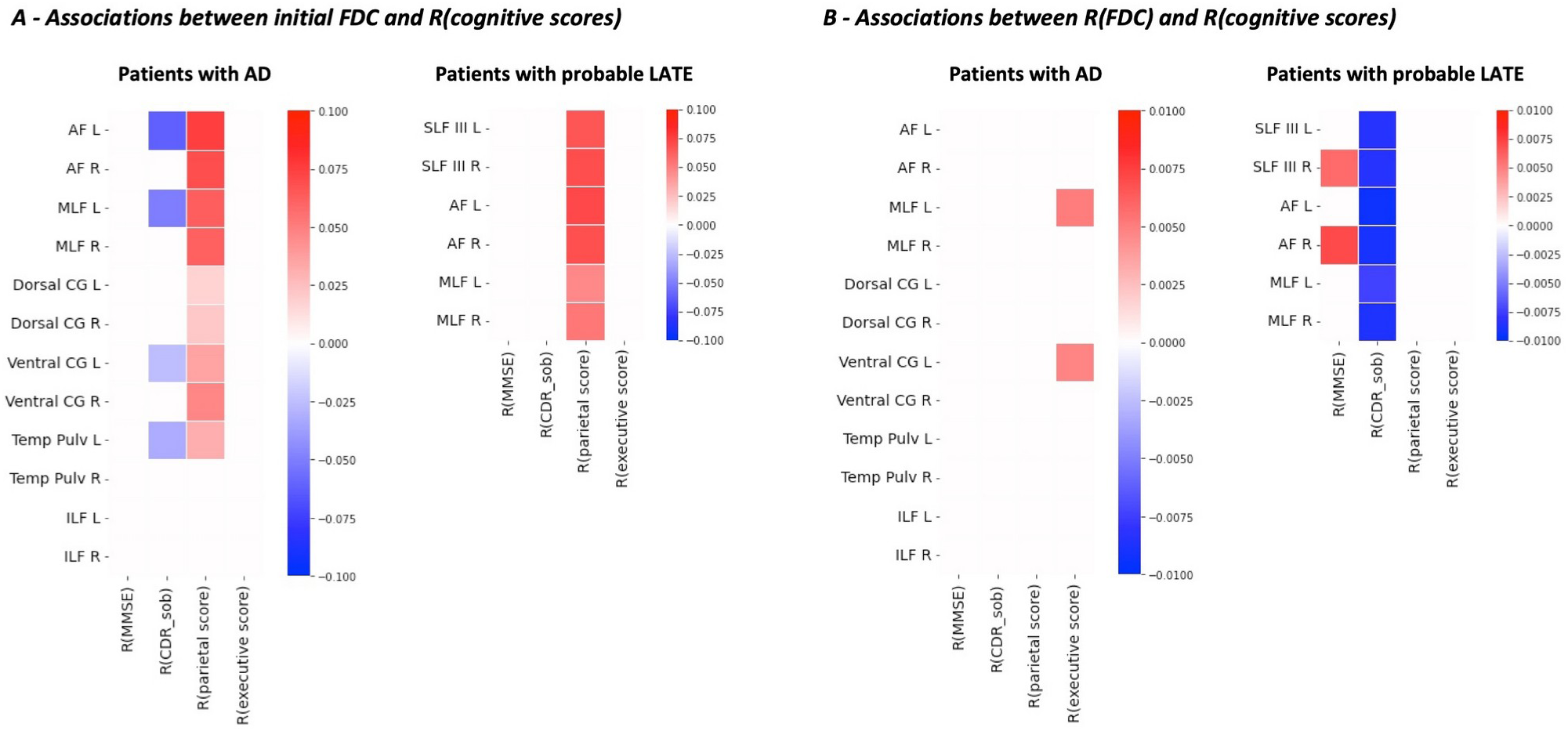
Associations between initial FDC values (A) or FDC rates of change (R(FDC)) (B) and cognitive/functional trajectories. Within each patient group and for each tract that showed a significant decline in FDC over time, we tested for associations between (A) initial FDC and cognitive/functional decline (R(MMSE), R(CDR_sob), R(parietal score), R(executive score)) using age, sex, and intracranial volume as covariates, and (B) FDC decline (R(FDC)) and cognitive/functional decline (R(MMSE), R(CDR_sob), R(parietal score), R(executive score)) using age and sex as covariates. Only significant associations (p < 0.05) are shown, with the color representing the strength of the association. Note that, contrary to the MMSE, parietal, and executive scores, the CDR_sob score increases with functional decline, hence resulting in opposite correlations. AF, arcuate fasciculus; CDR_sob, Clinical Dementia Rating sum of boxes; CG, cingulum; ILF; inferior longitudinal fasciculus; L, left; MLF, middle longitudinal fasciculus; MMSE: Mini Mental State Examination; R, right; SLF, superior longitudinal fasciculus; Temp Pulv, temporopulvinar bundle of Arnold.

Within the AD group, stronger FDC decline in the middle longitudinal fasciculus and ventral cingulum was associated with stronger decline in the executive composite score. Within the LATE group, we found that (i) stronger decline in FDC in the superior longitudinal fasciculi III, the middle longitudinal fasciculi and the arcuate fasciculi was associated with stronger functional decline; (ii) stronger decline in FDC in the right superior longitudinal III and arcuate fasciculi was associated with stronger decline in MMSE score (Figure 5B).

### Association between white matter alterations and PET imaging in the AD group

Higher amyloid-PET tracer uptake was associated with higher FDC values at baseline in the inferior longitudinal fasciculus (Figure 6A). We found no association with tau-PET imaging. Higher amyloid global cortical index was associated with stronger decline in FDC in the left ventral cingulum (Figure 6B).

**Figure 6.**
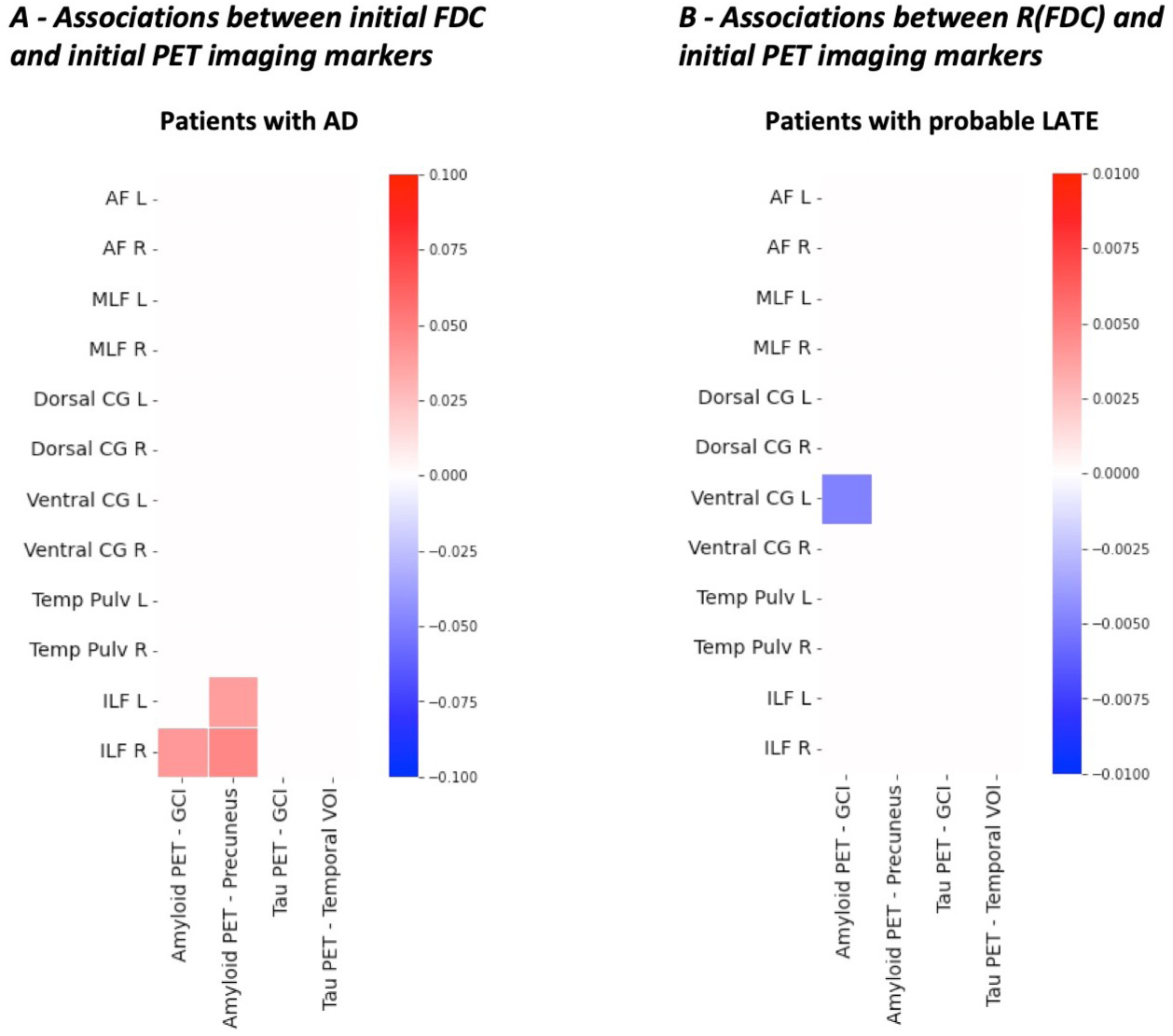
Association between initial FDC values (A) or FDC rates of change (R(FDC)) (B) and PET imaging markers at inclusion. In patients with AD, and for each tract that showed a significant decline in FDC over time, we tested for associations between (A) initial FDC and PET imaging markers at inclusion (amyloid GCI, amyloid SUVr in precunei, tau GCI, and tau SUVr in a temporal VOI) using age, sex, and intracranial volume as covariates, and (B) FDC decline (R(FDC)) and PET imaging markers at inclusion (amyloid GCI, amyloid SUVr in precunei, tau GCI, and tau SUVr in a temporal VOI) using age and sex as covariates. Only significant associations (p < 0.05) are shown, with the color representing the strength of the association. Please note that associations with amyloid-PET imaging markers were performed on n=15 patients with AD, as data were missing for one patient with AD, and that associations with tau-PET imaging markers were performed on n=14 patients with AD, as data were missing for two patients with AD. AF, arcuate fasciculus; CG, cingulum; GCI, global cortical index; ILF; inferior longitudinal fasciculus; L, left; MLF, middle longitudinal fasciculus; R, right; SLF, superior longitudinal fasciculus; SUVr, standardized uptake value ratio; Temp Pulv, temporopulvinar bundle of Arnold; VOI, volume-of-interest.

## Discussion

In this two-year longitudinal study, we investigated, for the first time to the best of our knowledge, the progression of damage to WM fiber bundles measured by fixel-based analyses in early AD and probable LATE. We found that WM fiber bundle alterations progressed significantly in both patient groups after two years in tracts connecting the temporal lobe to the parietal and frontal lobes, most of which were already significantly altered at baseline, such as the arcuate and middle longitudinal fasciculi. We also found a differential progression of WM damage according to pathology. Analysis of the associations between WM fiber bundle alterations at baseline and clinical progression suggested the involvement of alterations in the arcuate and middle longitudinal fasciculi in cognitive decline in AD and LATE, as well as of damage of the ventral and dorsal cingulum and temporopulvinar bundle of Arnold in AD, and of the superior longitudinal fasciculus in LATE.

In AD, the longitudinal trajectories of WM fiber bundle alterations have not been extensively studied and have mainly been investigated using the DTI model^10^, with one recent study using fixel-based analysis.^16^ Several studies reported widespread WM deterioration in the brain,^12,13,15^ while others described significant WM alterations in more specific tracts, such as the corpus callosum,^11,12,16,37,38^ the ventral cingulum,^11,13,16,38–41^ the fornix,^16,40^ tracts of the temporal lobe,^16,38^ posterior tracts,^14^ the uncinate fasciculus,^42^ and the superior and inferior longitudinal fasciculi.^11,12,14,16,39^ Apart from the difference in imaging method, these works differed from our study in a number of respects: (a) only clinical inclusion criteria were used, with exception of two studies in which CSF biomarkers were measured;^11,39^ (b) the follow-up period was shorter, with exception of two studies with a 2-year or more interval;^16,39^ (c) AD patients were probably at a slightly more advanced stage of the disease; (d) several studies did not include longitudinal data for healthy controls.^11,12,16,38,42^ Finally, the number of patients was only slightly higher than that of our cohort.

In our study, we found that the progression of WM fiber bundle alterations over two years was stronger in AD compared to controls in the ventral and dorsal cingulum and in the inferior longitudinal fasciculus, consistent with the literature.^10,16^ We also observed WM fiber bundle alterations over time in the arcuate and middle longitudinal fasciculi, and in the temporopulvinar bundle of Arnold. This latter tract, reported in our previous cross-sectional study,^27^ connects the anterior temporal lobe to the pulvinar and lateral dorsal nuclei of the thalamus,^43,44^ the latter of which is affected by neurofibrillary tangles at the same stage as the hippocampus.^45^ It has recently been described in a study assessing pulvino-temporal connectivity as being possibly involved in the lexical retrieval process of picture naming.^46^ The worsening of WM fiber bundle alterations in tracts of the temporal and limbic lobes, but also in those connecting the temporal lobe to the parietal and frontal lobes, is consistent with the known propagation of hyperphosphorylated tau pathology^47^ as well as with the pattern of progression of cognitive impairment in AD. Interestingly, the severity of damage in the arcuate and middle longitudinal fasciculi and in the cingulum at baseline was associated with more pronounced functional and cognitive decline, especially in the parietal cognitive score, consistent with the expected neurofunctional role of these tracts. This suggests that alterations of these bundles could anticipate or predict clinical evolution, in particular the extension of cognitive impairment to parietal functions in initially amnestic patients. We found fewer associations between the progression of WM fiber bundle alterations and cognitive decline, apart from relationships with decline in executive functions, the significance of which is uncertain. Our results highlight the importance of the middle longitudinal fasciculus, which connects the superior temporal gyrus and the temporal pole to the parietal lobe, in particular to the angular gyrus, and the consequence of its alteration on clinical decline in AD.

In LATE, no previous work has reported data on longitudinal alteration of WM, and only few recent studies have been conducted to explore WM alterations cross-sectionally. In patients with *post-mortem* confirmation, several studies have shown WM alterations in the temporal lobe^17-21^ and in temporo-temporal, temporo-basal ganglia, and temporo-frontal connections.^17,18^ In our cohort, we observed that the deterioration of WM fiber bundles over time was higher in LATE than in controls in the arcuate and middle longitudinal fasciculi, similarly to AD, and in the ventral section of the superior longitudinal fasciculus (SLF III), that connects the temporoparietal junction to the inferior frontal gyrus, in LATE only. These tracts were associated with cognitive decline, with the smaller magnitude of WM alterations compared to AD being congruent with slower cognitive decline, consistent with the literature. However, this patient group was smaller, so these associations should be treated with caution.

Taken together, these results support the hypothesis of a role of WM fiber bundle alterations in the progression of AD and LATE. However, the mechanisms that induce WM fiber bundle alterations, as well as their relationship with proteinopathies, are not yet clearly explained. Neuropathological studies reported early WM alterations in AD, defined by myelin degeneration and axonal loss.^5-7^ One explanation often proposed is a Wallerian degeneration mechanism secondary to tau pathology and neuronal loss in associative cortices.^6^ However, in the AD group, we found no association between baseline tau-PET imaging and the progression of WM fiber bundle alterations and only weak associations with amyloid-PET imaging. While this may be due to the small sample size, it should be emphasized that such associations have not yet been robustly described in the literature. Using different diffusion MRI metrics, some studies suggested associations between ventral cingulum alterations and tau-PET in representative cortical regions.^21,26,48,49^ In Jacobs et al,^50^ baseline mean diffusivity in the ventral cingulum predicted tau accumulation in the connected posterior cingulate cortex in amyloid-positive participants. Using cross-sectional fixel-based metrics, Vanderlinden et al^16^ also described significant associations between WM alterations in the ventral cingulum and tau accumulation in the connected posterior cingulate cortex and in the hippocampus. However, the analyses were performed combining patients and controls, which may have contributed to the positive results. Conversely, using cross-sectional fixel-based analysis, Dewenter et al^25^ could not find significant associations between WM alterations and tau-PET in cortical regions connected to the tested tracts, and reported even weaker associations when considering only amyloid-positive participants. This latter study also showed that small vessel disease may play a role in WM fiber bundle damage as measured by fixel-based analyses and must be taken into consideration. A possible increase in WM vascular lesions materialized by hyperintensities on T2 MRI over time was not found to any significant extent in our patients. Current evidence points to a direct role for WM fiber bundle alterations in clinical decline, but the exact nature of their relationship with proteinopathies will merit further investigation in studies involving longitudinal tau PET data in more regions-of-interest.

One strength of this study was the very precise definition of patient groups in terms of clinical phenotype and pathophysiological biomarkers. Patients with early AD had typical amnestic AD phenotype, positive CSF biomarkers, and positive amyloid and tau PET imaging. Patients with LATE were defined by a progressive episodic memory impairment mimicking early AD and negative AD pathophysiological biomarkers (CSF and molecular PET imaging) as recently proposed in the LATE clinical criteria.^4^ In addition, we checked for the absence of other clinical neurological diagnosis at baseline and after two years of follow-up, as well as the negativity of the second tau PET imaging when available. All control subjects were amyloid-negative, thereby excluding preclinical AD. Even if it inevitably limits the sample size, this longitudinal study was monocentric thereby reducing equipment-related variability, which could interfere with fixel-based analyses. Finally, we used the fixel-based analysis method, which is more specific than the more commonly used DTI method, because it allows fiber crossings’ representation and the derivation of metrics specific to each fiber population within each voxel.^22-24^ In addition, the registration steps were carefully handled, with a 2-step registration method, in order to reduce variability and increase sensitivity.^32^

The present study has several limitations. Its main weakness is the limited sample size, as already highlighted above, which probably limits the statistical power and the generalizability of the results obtained. This limitation is inherent to longitudinal multimodal imaging studies in symptomatic AD. The lack of neuropathological confirmation of LATE is another limitation of the present study, although we followed the recently published clinical criteria.^4^ In addition, the presence of TDP-43 co-pathology in patients with AD cannot be excluded.^1,3^

In conclusion, this study showed that WM fiber bundle alterations were detectable in AD and probable LATE at a mild cognitive impairment stage and that they worsened over two years in both patient groups, with slightly distinct patterns between groups. WM fiber bundle alterations appeared to be associated with clinical progression. This highlights the need to investigate WM tracts in the future to better understand their role in the mechanism and progression of these diseases.

## Data Availability

Due to the clinical nature of the data, the data analyzed in this study are not freely available but can be made available by the authors, upon reasonable request.

## Acknowledgment

We are grateful to the staff of the Centre de Neuroimagerie de Recherche (CENIR), Salpêtrière Hospital for patient management during MRI acquisition, and to the chemical/radiopharmaceutical and nursing staff of the Service Hospitalier Frederic Joliot for the synthesis of [^11^C]PIB and [^18^F]-Flortaucipir and for patient management. We are also indebted to AVID Radiopharmaceuticals, Inc. for their support in supplying the Flortaucipir precursor and chemistry production advice.

## Author contributions: CRediT

**Aurélie Lebrun:** Conceptualization, Investigation, Data curation, Methodology, Formal analysis, Visualization, Writing – original draft, Writing – review & editing; **Yann Leprince:** Supervision, Conceptualization, Investigation, Data Curation, Methodology, Writing – review & editing; **Pauline Olivieri:** Resources, Data curation, Writing – review & editing; **Martin Moussion:** Resources, Writing – review & editing; **Camile Noiray:** Resources, Writing – review & editing; **Michel Bottlaender:** Supervision, Funding acquisition, Project administration, Conceptualization, Investigation, Data curation, Writing – review & editing; **Marie Sarazin:** Supervision, Funding acquisition, Project administration, Conceptualization, Investiagtion, Data curation, Writing – review & editing; **Julien Lagarde:** Supervision, Funding acquisition, Project administration, Conceptualization, Investiagtion, Data curation, Writing – review & editing

## Supplementary Materials

Supplementary materials associated with this article can be found in the online version.

