## Supplementary file 1 for "Progression of fiber bundle damage in amnestic Alzheimer’s disease and LATE: a 2-year fixel-based study"

### Supplementary Information

#### MRI and PET acquisition and processing

**MRI acquisition parameters:**

For each subject and each session, diffusion-weighted images were acquired using a pulsed gradient spin echo single-shot echo planar imaging sequence with anteroposterior phase encoding (echo time (TE) = 77 ms; repetition time (TR) = 7 s; voxel size = 1.3 mm isotropic; acquisition matrix = 184 x 184; 110 axial slices; multiband acceleration = 2; generalized autocalibrating partial parallel acquisition acceleration = 2; partial Fourier factor = 0.625). Each diffusion acquisition comprised three shells of diffusion with b-values of 200, 1700, 4200 s/mm$^2$ and 60 directions per shell, and three volumes without diffusion weighting (b = 0 s/mm$^2$).

T1-weighted images were also acquired for each subject and each session with a magnetization-prepared rapid gradient echo sequence (MPRAGE; voxel size = 1 mm isotropic; TE = 2.15 ms; TR = 2.4 s; inversion time = 1 s; flip angle = 9°).

**MRI preprocessing:**

The quality of T1-weighted images was checked visually, guided by MRIQC.[1] Brain masks were then extracted using HDBET[2] and transformed to preprocessed diffusion-weighted images using the 'epi_reg' tool, which uses boundary-based registration[3] from FMRIB's Software Library (FSL, version 6.0.5).[4] Preprocessing of diffusion-weighted images included visual quality control, denoising,[5] and dynamic correction of susceptibility-induced distortion artefacts, eddy current induced distortions, and head motion, using the 'eddy_openmp' version[6] from FSL with replacement of outliers[7] and susceptibility-by-movement correction[8] options. Due to the lack of a $B_0$ field map or blip-down acquisitions, we used Synb0-DisCo[9] to estimate the susceptibility-induced distortion artefacts on each acquired b = 0 volume. Estimation of the displacement field was then performed using the 'topup' algorithm[10] from FSL and all three reconstituted pairs of b = 0 images. After eddy, we verified the quality of the preprocessed data using the 'eddyqc' tool.[11]

**PET processing:**

All corrections were incorporated in an iterative OSEM reconstruction and partial volume effect was corrected by directly modeling the detector spatial resolution properties in the image reconstruction algorithm.[12,13] Parametric images were created using BrainVisa software (http://brainvisa.info) on averaged images over 40–60 min after injection of [$^{11}$C]Pittsburg compound B and over 80–100 min after injection of [$^{18}$F]-Flortaucipir. Standardized Uptake Value ratio (SUVr) parametric images were obtained by dividing each voxel by the corresponding value found in the eroded (4mm) cerebellar gray matter.

#### Definition of cognitive outcomes

We defined the following composite scores:

- *Verbal episodic memory score*: sum of the scores obtained for the free and cued immediate and delayed recalls of the free and cued selective reminding test
- *Parietal score*: sum of the scores obtained for word naming, gestural praxis and copying of the Rey-Osterrieth complex figure
- *Executive score*: sum of the scores obtained for the forward and backward digit spans, letter fluency (2 min) and similarities subtest of the Wechsler Adult Intelligence Scale III.

We have previously shown that these cognitive components correlate significantly with MRI sulcal morphology parameters in corresponding neuroanatomical brain regions (memory and temporal cortex, parietal functions and parietal cortex, executive functions and frontal cortex).[14]
