## Supplementary file 2 for "Progression of fiber bundle damage in amnestic Alzheimer’s disease and LATE: a 2-year fixel-based study"

**Processing pipeline: MRtrix3 (version 3.0.3) commands applied for each subject for longitudinal fixel-based analysis (commands in grey are identical to those detailed in Mrtrix3 documentation*)**

*https://mrtrix.readthedocs.io/en/latest/fixel_based_analysis/mt_fibre_density_cross-section.html

```
cd ../{subject}/{session}
```

- **Selection voxels GM/WM for response function estimation:**

```
dwi2response dhollander data_file.nii.gz response_dhollander_wm.txt response_dhollander_gm.txt response_dhollander_csf.txt -fslgrad bvec_file bval_file -shells 0,200,1700,4200 -mask mask.nii.gz -voxels voxels.nii.gz
```

- **Selection voxels CSF for response function estimation:**

```
5ttgen fsl t1_file 5tt.mif

mrtransform t1_file -linear epi1t1_matrix.txt -inverse 5tt_coreg.mif

dwi2response msmt_5tt data_file.nii.gz 5tt_coreg.mif response_5tt_wm.txt response_5tt_gm.txt response_5tt_csf.txt -fslgrad bvec_file bval_file -shells 0,200,1700,4200 -mask mask.nii.gz -voxels msmt_5tt_voxels.nii.gz
```

- **Compute mean response function across all control subjects:**

```
cd ../..

responsemean {list_paths_response_dhollander_wm} group_average_response_wm.txt

responsemean {list_paths_response_dhollander_gm} group_average_response_gm.txt

responsemean {list_paths_response_5tt_csf} group_average_response_csf.txt
```

- **Compute FOD and apply normalization:**

```
cd /{subject}/{session}

dwi2fod msmt_csd -fslgrad bvec_file bval_file -shells 0,200,1700,4200 data_file.nii.gz group_average_response_wm.txt wmfod.mif group_average_response_gm.txt gm.mif group_average_response_csf.txt csf.mif -mask mask.nii.gz

mrconvert -coord 3 0 wmfod.mif - | mrcat csf.mif gm.mif - vf.mif
```

```
mtnormalise wmfod.mif wmfod-norm.mif gm.mif gm-norm.mif csf.mif csf-norm.mif
-mask mask.nii.gz
```

- **Create population FOD template:**

*We used the population template (wmfod_template.mif) previously computed in Lebrun et al, 2024. The preprocessing pipeline used in this publication is described in the Supplementary materials of that publication. A discussion on the choice of population template is available in Supplementary Materials file 1 of this publication. Please note that we optimized the regularization parameters for the creation of the template, using this final MRtrix3 command:*

```
population_template fod_input -mask_dir mask_input wmfod_template.mif -
voxel_size 1.3 -nl_update_smooth 0.75 -nl_disp_smooth 0.75 -nl_niter
10,10,10,10,10,10,10,10,10,10,10,10,10,10,10,10
```

- **Compute intra-subject averages:**

```
cd ../{subject}

mrregister {ses_V02}/wmfod-norm.mif {ses_V01}/wmfod_norm.mif -mask1
{ses_V02}/mask.nii.gz -mask2 {ses_V01}/mask.nii.gz -nl_warp_full
sesV022sesV01_warpfull.mif -transformed_midway
{ses-V02}/fod_in_midway_space.mif {ses-V01}/fod_in_midway_space.mif

mrmath {ses-V02}/fod_in_midway_space.mif {ses-V01}/fod_in_midway_space.mif
mean midway_space.mif

warpconvert sesV022sesV01_warpfull.mif warpfull2deformation
{ses-V02}/sesV022midwayspace_warp.mif -midway_space -from 1

warpconvert sesV022sesV01_warpfull.mif warpfull2deformation
{ses-V01}/sesV012midwayspace_warp.mif -midway_space -from 2
```

- **Transform masks to the intra-subject average and create intra-subject average mask:**

```
mrtransform {ses-V01}/mask.nii.gz -warp {ses-V01}/sesV012midwayspace_warp.mif
-interp nearest -nan -datatype bit {ses-V01}/mask_in_midway_space.nii.gz

mrtransform {ses-V02}/mask.nii.gz -warp {ses-V02}/sesV022midwayspace_warp.mif
-interp nearest -nan -datatype bit {ses-V02}/mask_in_midway_space.nii.gz

mrmath  {ses-V01}/mask_in_midway_space.nii.gz
{ses-V02}/mask_in_midway_space.nii.gz min mean_mask_in_midway_space.nii.gz
```

- **Register intra-subject average to the population template:**

```
mrregister midway_space.mif -mask1 mean_mask_in_midway_space.nii.gz
wmfod_template.mif -nl_warp midway_subject2template_warp.mif
midway_template2subject_warp.mif -nl_scale 0.25, 0.25, 0.5, 0.5, 1 -nl_lmax
0,2,2,4,4 -nl_niter 5000 nl_update_smooth 0.8 n_disp_smooth 0.8
```

- **For each session, compose deformation fields from subject to intra-subject average (step 1) and from intra-subject average to population template (step 2):**

```
transformcompose {ses-V01}/sesV012midwayspace.mif midway_subject2template.mif
{ses-V01}/subject2template_warp.mif
```

```
transformcompose {ses-V02}/sesV022midwayspace.mif midway_subject2template.mif
{ses-V02}/subject2tempate_warp.mif
```

- **Transform native FODs to the population template for each session of each subject:**

```
cd {session}
```

```
mrtransform wmfod-norm.mif -warp subject2template_warp.mif -reorient_fod no
{sesV01}/fod_in_template_space_NOT_REORIENTED.mif -nan
```

- **Create template mask and fixel mask on template:**

*We used the template mask (*template_mask_edited_thr.nii.gz*) and the fixel mask (*fixel_mask_0.1_edited*) previously computed in Lebrun et al, 2024. The preprocessing pipeline used in this publication is described in the Supplementary materials of that publication. Please note that we removed from the template mask areas where the confusion between WM and CSF compartments among subjects was too extensive, as represented in orange on the following image of one axial slice of the population template.*

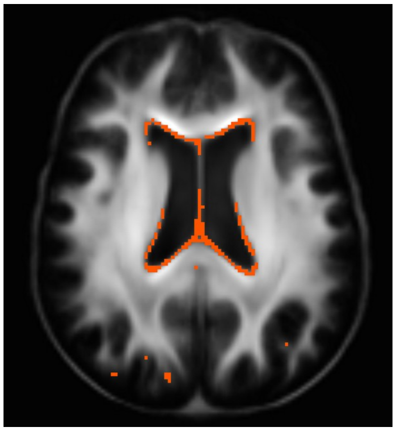

- **Estimate fixels for each session of each subject**

```
fod2fixel -mask ../template/template_mask_edited_thr.nii.gz
fod_in_template_space_NOT_REORIENTED.mif -fmls_peak_value 0.12
fixel_in_template_space_NOT_REORIENTED  -afd fd.mif
```

- **Reorient fixels for each session of each subject**

```
fixelreorient fixel_in_template_space_NOT_REORIENTED
subject2template_warp.mif fixel_in_template_space
```

- **Assign subject fixels of each session to template fixels**

```
fixelcorrespondence fixel_in_template_space/fd.mif
../template/fixel_mask_0.1_edited ../template/fd {subject}_{session}_fd.mif
```

- **Compute FC, log(FC) and FDC**

```
warp2metric subject2template_warp.mif  -fc
../template/fixel_mask_0.1_edited ../template/fc {subject}_{session}_fc.mif

cd ../template

mkdir log_fc

cp fc/index.mif fc/directions.mif log_fc

mrcalc fc/{subject}_{session}_fc.mif -log log_fc/{subject}_{session}_fc.mif

mkdir fdc

cp fdc/index.mif fdc/directions.mif fdc

mrcalc fd/{subject}_{session}_fd.mif fc/{subject}_{session}_fc.mif -mult
fdc/{subject}_{session}_fdc.mif
```

- **Compute rates of change of FD, log(FC) and FDC (R(FD), R(log(FC)), R(FDC))**

```
mkdir fd_diff

cp fd/index.mif fd/directions.mif fd_diff

mrcalc fd/{subject}_ses-V02_fd.mif fd/{subject}_ses-V01_fd.mif -subtract
{subject_time_interval_between_ses-V01_and_ses-V02} -divide
fd_diff/{subject}_diff_fd_ses-V02_ses-V01.mif

mkdir log_fc_diff

cp log_fc/index.mif log_fc/directions.mif log_fc_diff
```

```
mrcalc log_fc/{subject}_ses-V02_fd.mif log_fc/{subject}_ses-V01_fd.mif
-subtract {subject_time_interval_between_ses-V01_and_ses-V02} -divide
log_fc_diff/{subject}_diff_fc_ses-V02_ses-V01.mif

mkdir fdc_diff

cp fdc/index.mif fdc/directions.mif fdc_diff

mrcalc fdc/{subject}_ses-V02_fd.mif fdc/{subject}_ses-V01_fd.mif -subtract
{subject_time_interval_between_ses-V01_and_ses-V02} -divide
fdc_diff/{subject}_diff_fdc_ses-V02_ses-V01.mif
```

- **Compute whole brain tractography and filter tractogram**

```
tckgen -angle 22.5 -maxlen 250 -minlen 10 -power 1.0 wmfod_template.mif
-seed_image template_mask_edited_thr.nii.gz -mask template_mask.nii.gz
-select 20000000 -cutoff 0.1 tracks_20_million.tck

tcksift tracks_20_million.tck wmfod_template.mif tracks_2_million_sift.tck
-term_number 2000000
```

- **Generate fixel to fixel connectivity**

```
fixelconnectivity fixel_mask_0.1_edited/ tracks_2_million_sift.tck matrix/
```

- **Smooth fixel data using fixel to fixel connectivity**

```
fixelfilter fd_diff smooth fd_smooth_diff -matrix matrix/

fixelfilter log_fc_diff smooth log_fc_smooth_diff -matrix matrix/

fixelfilter fdc_diff smooth fdc_smooth_diff -matrix matrix/
```
