## Supplementary file 3 for "Progression of fiber bundle damage in amnestic Alzheimer’s disease and LATE: a 2-year fixel-based study"

### Supplementary Discussion – on the choice of template

In this study, we used as the common inter-subject template the one we previously computed (Lebrun et al, 2024) using the FODs obtained at session 1 of 8 patients with AD, 8 patients with probable LATE patients, and 16 healthy controls. This choice of template allowed us to reuse the same fiber bundle reconstructions as in our previous work. It results in a template computed using only images acquired at session 1, which makes it technically biased with respect to the analyzed images, because session 1 and session 2 could have different average registration distances to the template. However, we argue that this slight imbalance has a negligible effect on the results:

- The two-step registration method used to perform the longitudinal analysis greatly minimizes the influence of the registration distance from each time point to the template. Indeed, both time points of each subject use the same subject-to-template registration, with the difference between time points being based solely on an intra-subject registration, where the template does not come into play. Therefore, no bias in the intra-subject measurements can be introduced by a difference in registration distance to the template.
- The two-year time interval between sessions is small with respect to the age dispersion in the cohort (standard deviation ranging from 3.5 to 6.0 years within each group see Table 1). Any between-group difference in registration distance is likely to be driven by age differences rather than the choice of template, and that effect is accounted for by including age as a covariate.
- We have successfully reproduced a part of the presented results using a fully unbiased template, with essentially identical results, as part of  a separate study in which we explored the impact of the two-step registration method in longitudinal fixel-based analyses (Lebrun et al, 2025). In that study, we included the same patients with AD and healthy controls but we did not include patients with LATE. We computed an unbiased longitudinal template by using the intra-subject averages of each selected subject to compute the template, instead of using the FODs obtained at session 1. We then performed the same within-group whole-brain fixel-based analysis within the AD group as the one we performed in the present study (see Figure 1), to investigate the decrease in fixel metrics over two years at the level of each fixel. The results obtained in each study are the following:

***Results presented in Lebrun et al, 2025, obtained with an unbiased longitudinal template***

***Results presented in the present study, with a template computed from images acquired at session 1***

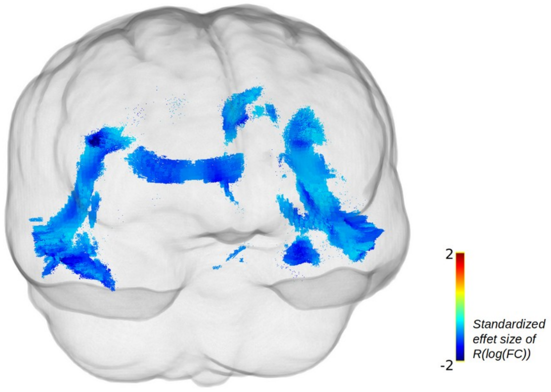
