## Supplementary file 4 for "Progression of fiber bundle damage in amnestic Alzheimer’s disease and LATE: a 2-year fixel-based study"

### Supplementary Table - Tract extremity regions

| Bundle name | Extremity A | Extremity B |
|---|---|---|
| Callosal fibers – Superior Frontal gyri | Truncated superior frontal gyrus | Contralateral truncated superior frontal gyrus |
| Callosal fibers – Caudal middle frontal gyri | Caudal middle frontal gyrus | Contralateral caudal middle frontal gyrus |
| Callosal fibers – Precentral gyri | Precentral gyrus | Contralteral precentral gyrus |
| Callosal fibers – Precunei | Precuneus | Contralateral precuneus |
| Cerebello-thalamo-cortical tract | Cerebellum | Contralateral truncated superior frontal gyrus (+ thalamus as included region) |
| Superior Longitudinal Fasciculus III | TractSeg beginning mask | TractSeg ending mask |
| Arcuate Fasciculus | TractSeg beginning mask | TractSeg ending mask |
| Middle Longitudinal Fasciculus | TractSeg beginning mask | TractSeg ending mask |
| Dorsal Cingulum | Caudal + rostral anterior cingulate | Isthmus cingulate + Precuneus |
| Ventral Cingulum | Hippocampus + temporal pole + entorhinal cortex | Isthmus cingulate + Precuneus |
| Temporopulvinar Bundle of Arnold | Temporal pole | Thalamus |
| Inferior Longitudinal Fasciculus | TractSeg beginning mask | TractSeg ending mask |
| Uncinate Fasciculus | TractSeg beginning mask | TractSeg ending mask |

Tractography was also performed using appropriate exclusion masks to remove aberrant fibres. The masks of the named regions were computed as the FreeSurfer probabilistic maps of presence in the common FOD template space (with the Desikan-Killiany cortex parcellation).

The cerebello-thalamo-cortical tract was subsequently cut in inferior and superior parts with to respect to the thalamus.
